# Identification of novel associations of candidate loci with Alzheimer’s disease by leveraging the shared genetic basis with hippocampal volume

**DOI:** 10.1101/2024.10.01.24314738

**Authors:** Chenyang Jiang, Sven J. van der Lee, Niccolo Tesi, Wiesje M. van der Flier, Betty M. Tijms, Lianne M. Reus

## Abstract

**Objective:** Alzheimer’s disease (AD) is a complex neurodegenerative disorder with a considerable genetic contribution that remains not fully understood. The hippocampus plays a critical role in learning and memory, with its volume loss being a core hallmark of AD. Hippocampal volume also has a strong heritable component and its genetic underpinnings may help us to understand the complex biological mechanism underlying AD.

**Methods:** We performed cross-trait analysis of exisiting GWAS data on late-onset AD and Hippocampal volumes using the conjunctional false discovery rate (conjFDR) framework to identify the specific shared genetic basis.For identified SNPs, we performed functional annotation and phenome-wide association studies (PheWAS).

**Results:** Our cross-trait analyses identified 11 non-APOE lead genetic loci, of which 7 loci showed discordant directional effects (loci associated with decreased risk for AD and smaller hippocampal volumes, and vice versa). We found that *SHARPIN* and *TNIP1* genes play a role in AD by affecting the hippocampal volumes. In addition, we observed 9 novel AD-hippocampus loci in genes previously implicated in AD (*IGIP* and *ACE*) and novel AD-genes (*KCTD13, HINT1, SH3TC2, FAM53B, TPM1, IL34* and *SSH2*). Phenome-wide association study highlighted varying degrees of pleiotropy, including brain imaging measurements, white blood cell markers, red blood cell markers, and lipids in multiple shared loci.

**Conclusions:** Our integrating GWAS study reveals a shared genetic basis between AD and hippocampal volumes. By integrating GWAS summary statistics for these two traits, we identified both novel and previously reported AD-hippocampus loci. Functional analysis highlights the roles of immune cells and lipid markers in the shared loci and traits, suggesting a shared neurobiological basis for both traits.

## Introduction

Alzheimer’s disease (AD) is a progressive and irreversible brain disorder affecting over 50 million people worldwide(1). Genome-wide association studies (GWAS) have identified over 80 genetic variants associated with AD, but together these explain only a small fraction of its high heritability(2). This discrepancy may be due in part to the common practice of using clinical AD as a single outcome in GWAS studies, overlooking the fact that AD is a heterogeneous condition. Greater genetic variance could potentially be captured by integrating GWAS data from multiple phenotypes, which could help to guide the development of targeted treatments and strategies.

A core symptom of AD is memory impairment, which is closely related to brain atrophy. The Hippocampal volumes can be assessed using structural magnetic resonance imaging (MRI)(3)(4), and previous GWAS studies have indicated that this measure is highly heritable (17% heritability)(5). Prior studies in young adults suggested that the genetic markers for AD confer life-long susceptibility to different brain structure: both the major genetic risk factor for AD Apolipoprotein E (APOE)-e4, and APOE-excluded polygenic risk score (PRS) were shown to have independent effects on hippocampal volumes(6–9). These observations suggest that the genetic architecture of AD overlaps with genetic variations that influence individual variability in hippocampal volumes.

One way to study shared genetic architecture between two traits is through conjunctional false discovery rate (conjFDR) analysis, which aims to enhance the identification of shared individual genetic variants by leveraging the collective power of two GWAS studies(10). Previous studies using this approach in psychiatric disorders indicated that this could leverage auxiliary genetic information and brain morphology (10–12), leading to the identification of novel loci (13). We hypothesized that combining genetic data of AD and hippocampal volume, one of the core affected brain signature in AD, could improve the detection of novel AD-associated genetic markers.

In this study, we examined this hypothesis on the identification of shared genetic risk loci, by conducting a conjFDR analysis on the most recent GWAS on AD and bilateral hippocampal volumes (5, 14). To identify candidate genes with functional relevance, we then performed the downstream analysis of the discovered loci in Genotype-Tissue Expression (GTEx) data and other databases. The results of this study may elucidate pleiotropic effects on AD and hippocampal volumes that offer insights into causal neuropathological pathways underlying both traits.

## Methods

### 1. GWAS summary statistics

GWAS summary statistics were obtained from GWAS catalog [https://www.ebi.ac.uk/gwas/] for the AD GWAS (7) and Oxford Brain Imaging Genetics Server [https://open.win.ox.ac.uk/ukbiobank/big40/] for Imaging Phenotypes GWAS (5). Descriptive statistics for the GWASs used in this study are provided in Supplementary Table 1. Details for the genotyping procedure, quality control, and GWAS analysis are provided in the primary manuscripts for the AD GWAS and the hippocampal volumes GWAS. The AD GWAS dataset consisted of summary statistics including association P values, beta coefficients, standard errors, and effect alleles. This study included 85,934 clinically diagnosed/proxy AD cases and 401,577 controls, and found 75 AD-related risk loci at stage 1 analysis. The hippocampal volumes GWAS dataset consisted of summary statistics including association P values, beta coefficients, standard errors, and effect alleles based on 39,691 participants of the UK Biobank(15). Quality control (included sanity check and duplication removing) of the GWAS summary statistics was conducted in the publicly available toolbox GWASlab [https://github.com/Cloufield/gwaslab]. Coordinates of SNPs in the AD GWAS were lifted from hg38 to hg19 to align with the default genome assembly of the conjFDR software. We removed four regions with complex Linkage Disequilibrium (LD) patterns, including major histocompatibility complex region: 6:25119106–33854733; 8p23.1: 8:7200000-12500000; *MAPT* region: 17:40000000–47000000; and the *APOE* region: 19:45111942 – 45711941. A total of 12,025,887 SNPs were common in the two GWAS that were used for further analysis.

**Table 1.**
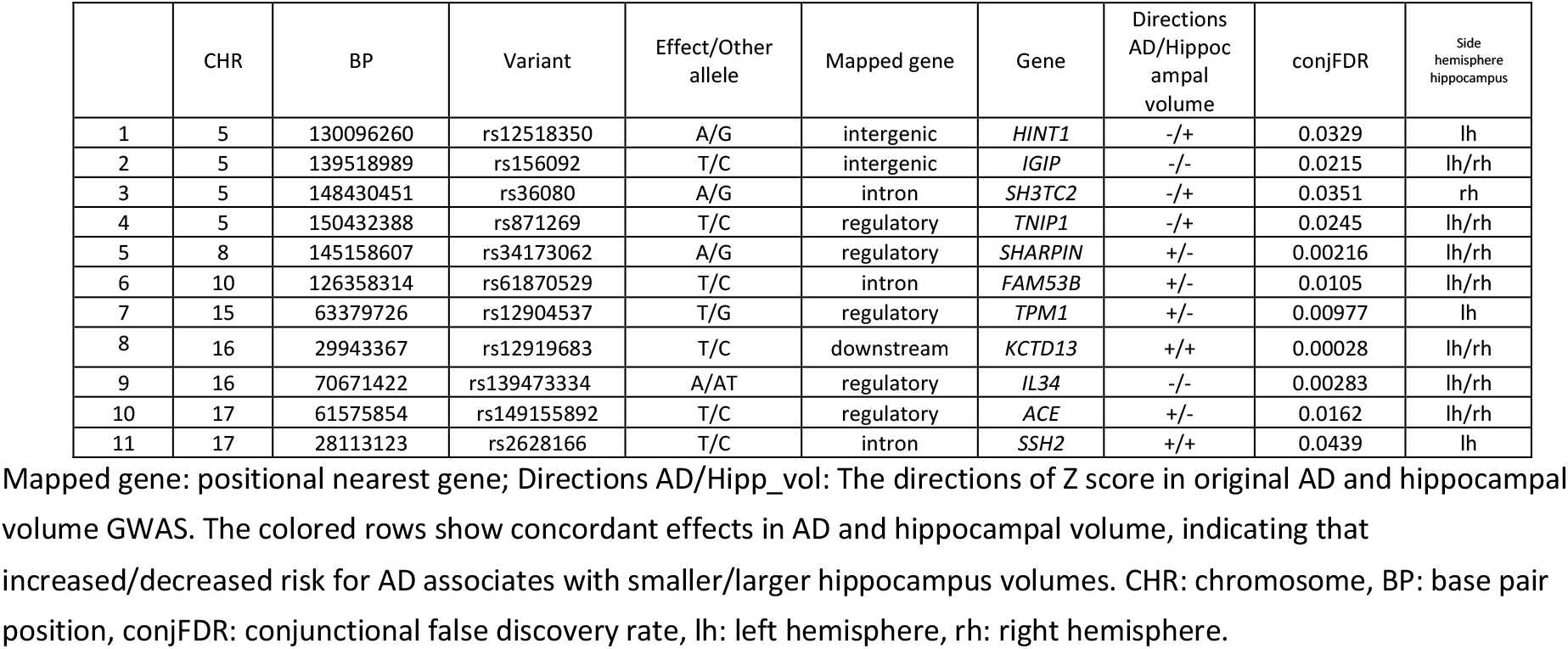
Common lead loci shared with Alzheimer’s disease and bilateral hippocampal volumes.

### 2. Shared Locus discovery

We applied conjFDR analysis to explore the shared genetic variants across the AD and hippocampal volume GWAS(10). The conjFDR analysis is built on an empirical Bayesian statistical framework and it leverages cross-trait SNP enrichment to improve the power to discover variants with a shared effect. We first used SNP associations of the hippocampal volumetric phenotype to re-rank test statistics and re-calculate the significance of associations between these SNPs and AD. Then, we inverted the roles of two phenotypes to re-calculate the significance of SNP associations to the brain volumetric phenotypes. The conjFDR analysis conservatively estimates the posterior probability that a SNP has no association with either trait, represented as a conjFDR value. This method can effectively boost the GWAS discovery power by leveraging auxiliary genetic information contained in a highly phenotypic-related trait. SNPs with a significance of conjFDR < 0.05 and having LD r2 < 0.1 within 1000 kb were defined as independent significant signals with ‘–clump’ function in PLINK(16).

### 3. Co-localization

To examine whether shared loci identified in the conjFDR analysis could be causal to both AD and (bilateral) hippocampal volume, we performed colocalization analysis using coloc(17) for all shared loci. Briefly, coloc uses a Bayesian approach to estimate the posterior probability (PP) that a given genetic locus is not associated with AD or hippocampal volume (hypothesis 0 [H0]), is uniquely associated with AD (H1) or hippocampal volume (H2), harbors two independent causal variants for each phenotype (H3), or harbors a shared causal variant for both AD and hippocampal volume (H4). Coloc anlaysis evaluated evidence of colocalization within a 500 kb region centered at each shared risk locus. The posterior probabilities for H4, i.e there is a similar causal locus for both AD and hippocampal volume, were used to evaluate the evidence for colocalization of shared loci.

### 4. Functional interpretation of candidate shared genetic variants

To study the potential functional consequences of loci shared across AD and hippocampal volumes, we used the functional annotation section of the snpXplorer (18) web server with default settings. This tool performs variant-to-gene mapping integrating variant consequences based on CADD score (19), and quantitative-trait-loci analyses (expression-QTLs and splicing-QTLs). Given the large number of variants associated with gene expression, we further performed colocalization analyses between shared loci and eQTL signals from GTEx database (20) to identify the possible candidate genes. To further characterize the significant shared loci, we conducted phenome-wide association studies (PheWAS). Briefly, PheGWAS is a powerful tool for exploring the associations between genetic markers and a wide range of traits, and can provide insights into the complex relationships between genetics and phenotypic traits. The traits that are associated with the list of SNPs with P < 1 × 10 –5 in all GWAS harmonized summary statistics in the MRC IEU OpenGWAS data infrastructure were extracted by using ‘phewas’ function of the R-package ‘ieugwasr’ (21). We performed quality control on the PheWAS results, which included standardizing trait names, filtering out traits with a sample size of less than 50,000 due to their low statistical power, and removing duplicated SNP-trait associations. To identify key pleiotropy, we selected the top three traits for each shared locus based on P-values for visualization.

## Results

### 1. conjFDR analysis identifies 11 genetic loci shared across Alzheimer’s disease and bilateral hippocampal volumes

We applied conjFDR analysis to identify shared genetic variants across AD and bilateral hippocampal volumes. To provide a visual pattern of pleiotropic enrichment between the two phenotypes, we generated conditional quantile–quantile (Q–Q) plots (Figure 1) from the conjFDR results. The conditional Q–Q plots showed a strong enrichment pattern for AD given hippocampal volumes, and vice versa. SNPs with a significance of conjFDR < 0.05 and LD r2 < 0.1 within 1000 kb were defined as independent significant signals. We identified 10 lead loci shared by AD and left hippocampal volume, and 8 common loci shared by AD and right hippocampal volume (Supplementary Table 2). After merging lead signals for left and right hippocampal volumes with similar LD patterns (i.e., LD blocks closer than 250 kb), we identified 11 unique lead loci shared by AD and hippocampal volumes (see Table 1 for a list of all shared loci). We then compared the effect direction on AD risk and hippocampal volumes. There were 7 loci with opposite direction of effects (increased AD risk coincided with smaller hippocampal volumes), and 4 loci shows same direction (Figure 2).

**Figure 1.**
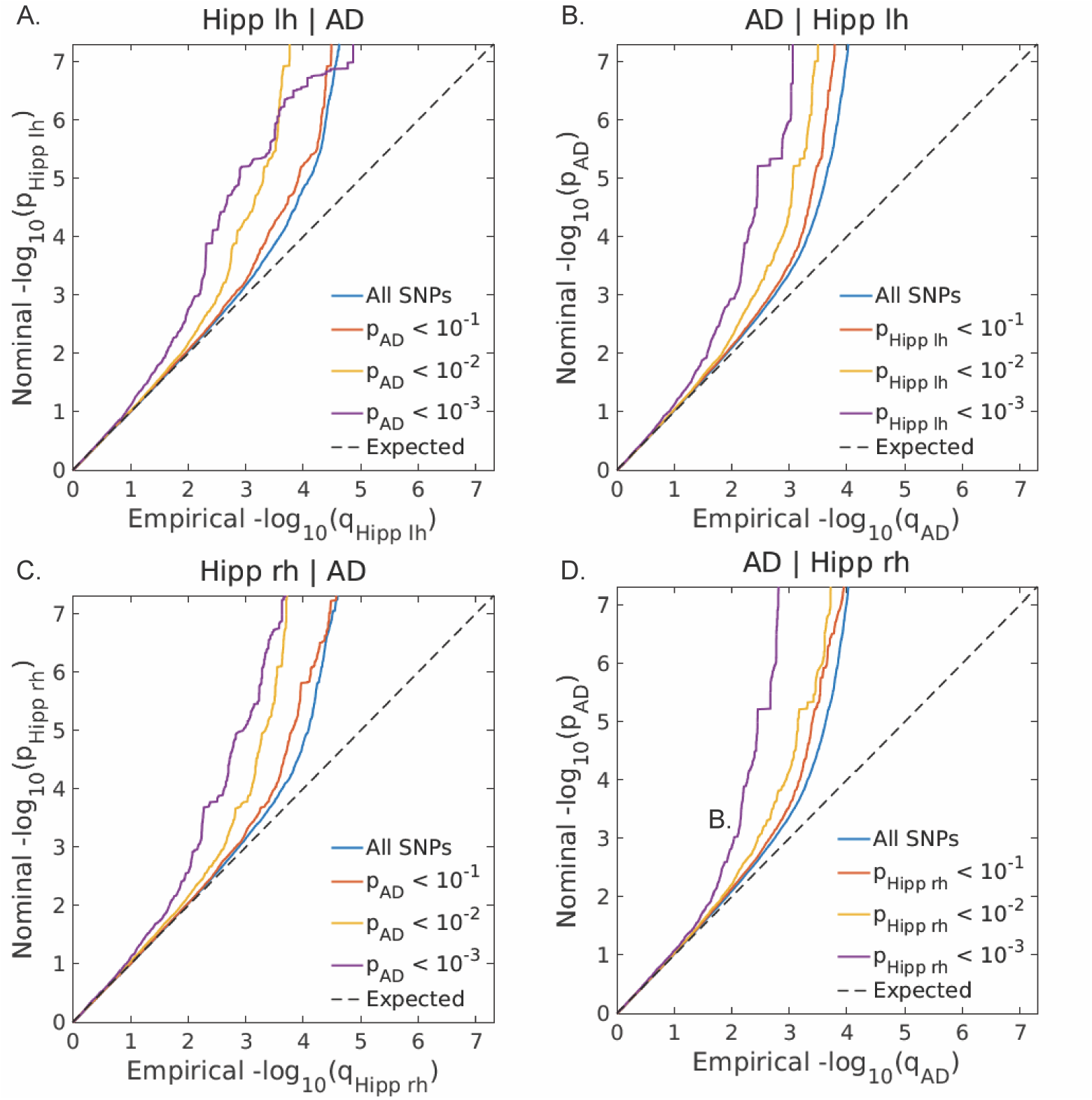
Conditional quantile–quantile plots. Conditional Q–Q plots of nominal versus empirical AD−log10p values below the standard GWAS threshold of P<5×10^−8^ as a function of significance of association with bilateral hippocampal volumes, at the level of P≤ 0.1, P≤ 0.01, P≤ 0.001, respectively (left panel). Conditional Q–Q plots of nominal versus empirical bilateral hippocampal volumes−log10p values below the standard GWAS threshold of P<5×10^−8^ as a function of significance of association with AD, at the level of P≤ 0.1, P≤ 0.01, P≤ 0.001, respectively (right panel). The dashed lines indicate the null hypothesis. The blue line shows association P values in the main trait of interest (AD) for all SNPs irrespective of their association P values in hippocampal volumes. Red, yellow, and purple lines show association P values in the main trait for successively nested subsets of SNPs with increasing significance in the conditional trait. The consistently increasing degree of leftward deflection for subsets of variants with higher significance in the conditional trait in both directions indicates substantial polygenic overlap between AD and hippocampal volumes.

**Figure 2.**
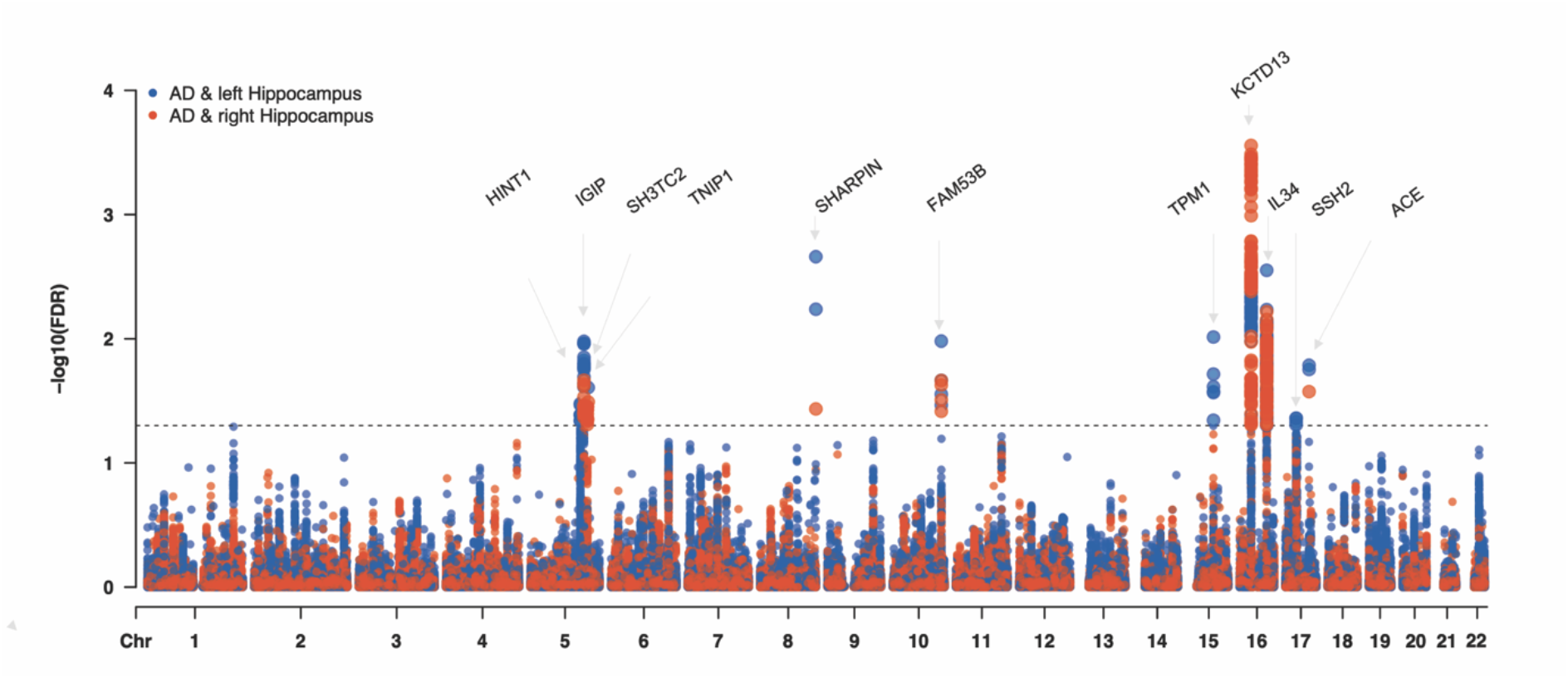
Common genetic variants jointly associated with Alzheimer’s disease and bilateral hippocampal volumes. The x-axis stands for the base pair position and y-axis represents -log10 transformed conjunctional false discovery rate values with a black horizontal line reflecting significance. The lead signals which LD blocks closer than 250 KB associated the left and right hippocampal volumes will be merged into one locus. KB: Kilobyte; Chr: chromosome; FDR: false discovery rate; AD: Alzheimer’s disease; conjFDR: conjunctional false discovery rate; LD: linkage disequilibrium

### 2. Shared Alzheimer’s disease-hippocampus loci act as quantitative trait loci (QTLs)

To gain more insight into the potential functional consequences of the 11 lead SNPs for AD and hippocampal volumes, we performed positional, eQTL, and sQTL mapping using existing gene expression databases (18) (20). A full overview of the functional annotation of all 11 shared loci is presented in Supplementary Table 3. The majority of the signals were regulatory (45%), intronic (27%) and intergenic (18%). Three lead loci had supportive evidence from colocalization analysis for a shared causal genetic variant between AD and hippocampal volume, including *TNIP1, SHARPIN* and *IGIP* (range posterior probability PP-H4 =0.74-0.99, Supplementary Table 4). The lead variant located on an intron of *TNIP1* (TNFAIP3 Interacting Protein 1) (rs871269; conjFDR=0.025) (Supplementary Figure 3) associated with gene expression of *LACTB, RAB8B* and *RPS27L* of multiple brain tissues. This potential protective variant for AD reached suggestive genome-wide significance in the Bellenguez study stage1 meta results (P=3.7×10-5), and genome-wide significance in the stage1 and stage2 meta-analysis (P=8.7×10-9)(14), demonstrating the potential for enhancing gene discovery power by integrating hippocampal volumes in this study. The lead variant located on an exon of *SHARPIN* (rs34173062; conjFDR=0.0245) (Figure 3a) is missense variant is a known AD risk locus and is involved in postsynaptic function (22). This variant acts as eQTL, with trans effects of rs34173062 [effect allele A] on increased gene *ZNF707* expression in the anterior cingulate. The lead variant rs156092 (conjFDR=0.0215) is located on the previously reported AD risk gene *IGIP*, but this specific locus is new to AD. No QTL effects have been identified for this genetic variant. Another shared locus is rs12919683 (conjFDR=0.00028), which is located on *KCTD13* (Figure 3c). This locus seems to have multiple trans QTL effects, including effects on brain gene expression of *YPEL3, MAPK3, GDPD3, TBX6, INO80E, ASPHD1* and *BOLA2*, and sQTL effects *DOC2A* splicing in multiple brain tissues.

**Figure 3.**
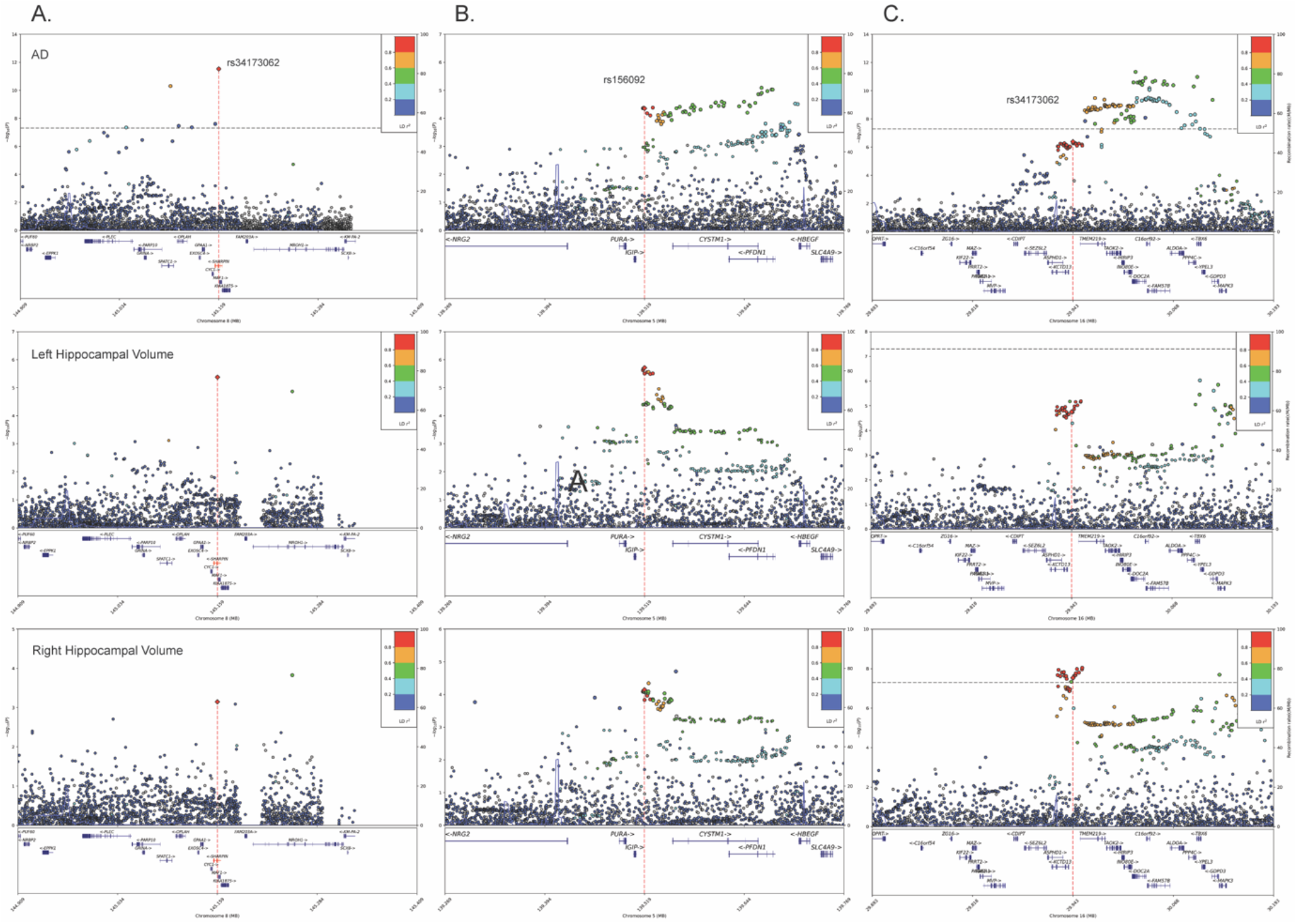
Regional plots: Single-nucleotide polymorphism-level associations with AD, bilateral hippocampal volumes at *SHARPIN, IGIP* and *KCTD13* locus, displayed as a locus zoom plot. The index SNP is annotated in purple, and nearby SNPs within 500KB ranges are colored according to how strong they are in LD (r2) with the index SNP, as derived from the 1000 Genomes LD reference. a: Index SNP rs34173062; b: Index SNP rs156092; c: Index SNP rs12919683. AD: Alzheimer’s disease; KB:Kilobyte ; LD: linkage disequilibrium

### 3. Functional interpretation based on Phenome-wide association studies

By systematically analyzing all significant associations (P < 1 × 10 –5) in the MRC IEU OpenGWAS database, in total we identified 442 associations for the 11 significant shared loci. Most of the associations were found in *SSH2* (30.0%), *KCTD13* (24.4%), and *SHARPIN* (13.8%) locus, (Supplementary Table 5). Among all the associated traits, we found half to be brain neuroimaging measurements (51.5%), 21.5% are classified to the body measurements category, which is a collection of 57 traits in the database, and 7.2% of traits belong to white blood cell category (Supplementary Table 5). For selecting key pleiotropic traits (Figure 4), we identified the strongest associations for each locus. We observed 5 loci including *HINT1, SHARPIN, SSH2, FAM53B*, and *TNIP1* related to white blood cell markers, and KCTD13 showed the strongest associations with red blood cell markers.

**Figure 4.**
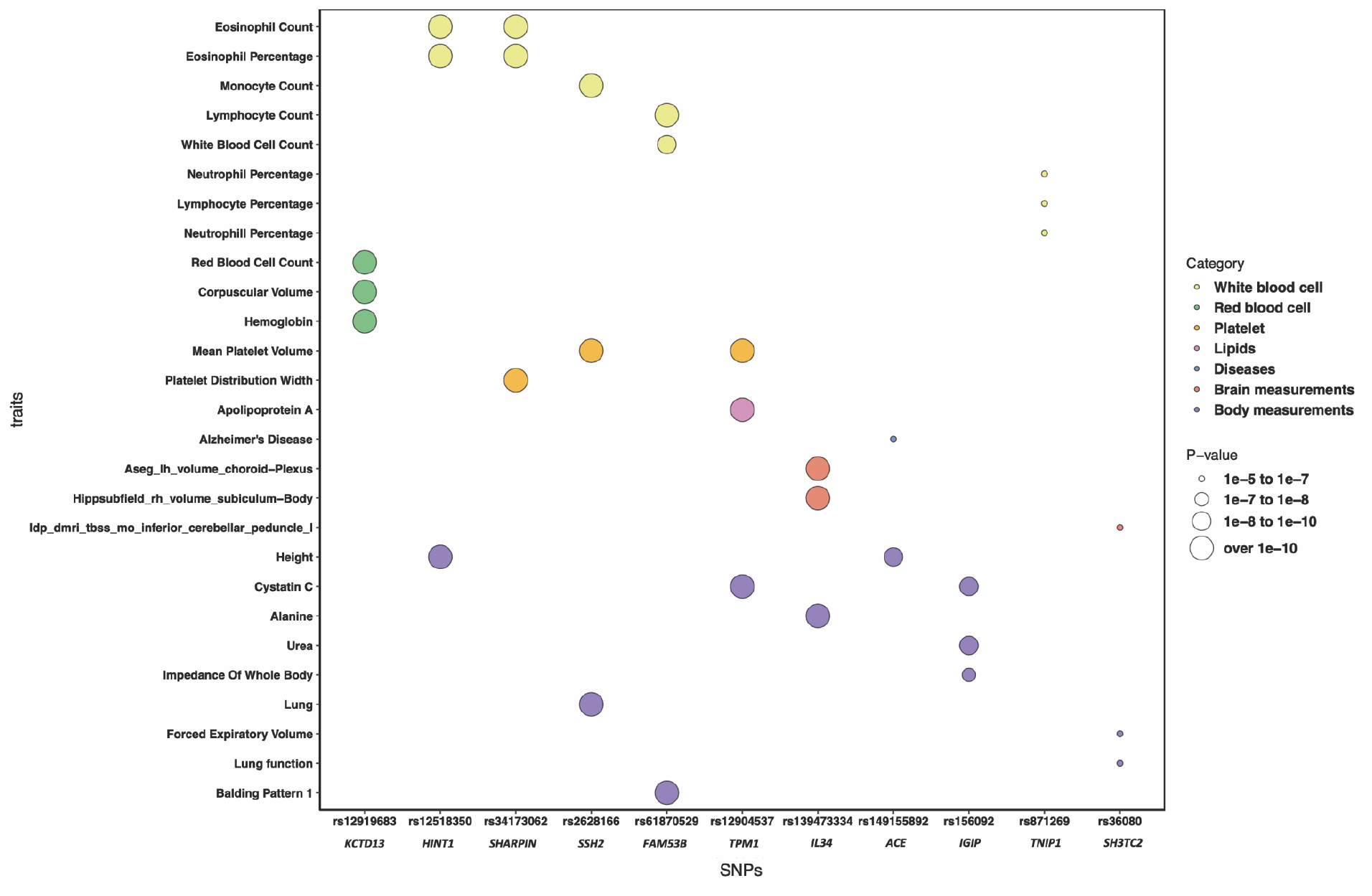
Functional interpretation based on Phenome-wide association studies. Top hits from the Phenome-wide association studies (PheWAS) were identified by searching for key pleiotropic traits associated with a list of SNPs with P < 1 × 10^−5^ in the MRC IEU OpenGWAS data. The x-axis indicates shared loci, while the y-axis represents related traits from the PheWAS analysis. The circle size corresponds to the p-values, and the colors represent different trait categories. The related traits belong to the following categories: white blood cell, red blood cell, platelet, lipids, diseases, brain measurements, and body measurements.

## Discussion

In this study, we aimed to identify the shared genetic architecture underlying AD and hippocampal volumes. By leveraging the collective power of GWAS studies on these two traits, we were able to boost AD gene discovery and identify 11 loci that were associated with both AD and hippocampal volumes. Shared loci included two previously identified (*TNIP1* and *SHARPIN*) (14, 23) and 9 potential novel risk AD risk loci located on *KCTD13, IGIP, HINT1, SH3TC2, FAM53B, TPM1, IL34, ACE* and *SSH2*.

We identified two known AD loci to be associated with hippocampal volume, one of them including *TNIP1* (rs871269) located on chromosome 5(14). *TNIP1* has been associated with hippocampal sclerosis and amyotrophic lateral sclerosis (ALS) (24). Its product, *TNIP1*, has an inflammatory function, but it has also been expressed in hippocampal and cerebellar areas during the development of the central nervous system, highlighting its crucial role in the development of brain (25). Previous studies have indicated that the causal genetic variant for AD and ALS underlying *TNIP1* may be the nearby-located gene *GPX3* (26). Accordingly, one recent study reported that *TNIP1* (rs34294852) affects levels of CSF GPX3, a protein that protects against oxidative stress, as AD progresses (26). Here, we report that *TNIP1* also may influence gene expression of *LACTB, RAB8B* and *RPS27L*, of which the latter two are both involved in autophagy (27, 28). And recent community study found cortical protein, *LACTB* is implicated in Alzheimer dementia (29). Further research is needed to understand through which specific immune-related mechanisms, the *TNIP1* locus both influences hippocampal volume and risk for AD. The second known AD locus, rs34173062 on *SHARPIN*, has been associated with late-onset AD in various ethnic groups (30). This missense variant is involved in postsynaptic function (22). Another GWAS study showed that the medial temporal circuit could be used as another imaging phenotype and the SNP rs34173062 on the *SHARPIN* gene had a genetic modifying effect on its atrophy (31). Those shared variants may play a role in AD pathogenesis by affecting the hippocampal volumes, which is instrumental in refining their interrelationships.

Additionally, we have identified 9 potential novel AD loci. One intriguing shared signal was *KCTD13*, which is located on the 16p11.2 locus. *KCTD13* belongs to the KCTD family, of which many members are associated with neuropsychiatric disorders (32). *KCTD13* has been documented to act as a cullin-3 adaptor for the ubiquitination and degradation of RhoA, a crucial small GTPase protein regulating the actin cytoskeleton, pivotal for neuronal development and synaptic function. Deletion of the entire *KCTD13* gene results in elevated RhoA expression, loss of dendritic spines, and diminished synaptic activity in hippocampus CA1 region (33). RNA-seq analyses of gene expression profiles from the cortex and hippocampus of KCTD13-deficient (exon 2-deleted) mice unveiled altered signaling pathways critical for neurodevelopment, including synaptic formation and reduced spine density in the hippocampus (33, 34). Our PheWAS results revealed the main role of *KCTD13* locus in red blood cell marker, including cell count, corpuscular volume and hemoglobin. Previous study has found its the U-shaped associations with incident dementia risk, and hemoglobin and anemia were associated with brain structure alterations (35). All those findings raise the possibility that genetically-induced changes in blood cell markers may affect AD risk, via structural neural changes in brain.

Of the 11 genetic variants that were shared between AD and hippocampal volume, 7 signals had discordant effect directions, indicating that the minor allele was associated with increased risk for AD and smaller hippocampal volumes (and vice versa, where decreased risk for AD associates with larger hippocampal volumes). This association between smaller hippocampal volumes at middle-age and increased risk for AD is in line with brain capacity theory (36, 37). According to this theory, grey matter volume is a potential quantitative brain reserve, indicating that larger brain volumes protect against the consequences of neuronal degeneration (38). The 4 loci with concordant effects (i.e., increased AD risk and larger hippocampal volumes) imply more complicated neurodevelopmental process in the life stage. For instance, larger hippocampal volumes have been found to correlate with autism and children with fragile X syndrome (39), and these neurodevelopmental conditions are believed to share genetic and mechanistic overlaps with AD (40). The left and right hippocampal volumes did not show the exact same genetic overlap with AD: three loci were specific to left hippocampal volumes, and one locus was only associated with right hippocampal volume. Subcortical volume asymmetries are considered an important aspect of brain organization and play an important role in AD progression, and multiple genetic variants have been identified with brain asymmetries (41, 42). Furthermore, we observed that several loci only associated with hippocampal volume of a single hemisphere: three loci were specific to the left hippocampal volume, and one locus was only associated with the right hippocampal volume.

Our study has certain limitations. Firstly, there may be overlap in the samples used across the GWAS in our cross-trait analysis, which could lead to overestimating the effects of shared signals. While we used the largest GWAS summary statistics available for our current analyses, independent data is needed to validate identified shared loci in the future. Second, while our research provides genetic evidence for a shared role of immune cell and lipids markers in AD and hippocampal volumes, the specific pathway involved in this process needs experimental validation.

In summary, our study integrated GWAS summary statistics on AD and hippocampal volumes to uncover the shared genetic architecture between these traits. We identified two known AD loci located in *SHARPIN* and *TNIP1*, along with nine novel risk loci, including *KCTD13* as shared genetic variants. Through gene annotation and QTL analysis, we pinpointed the potential mapped genes for these shared loci, most of which are protein-coding. Additionally, we highlighted the significant roles of immune cells and lipid markers as shared mechanisms in the gene-hippocampus-AD pathway.

## Supporting information

It contains required supplemenatry figures and tables.

## Data Availability

GWAS summary statistics were obtained from GWAS catalog [https://www.ebi.ac.uk/gwas/] for the AD GWAS and Oxford Brain Imaging Genetics Server [https://open.win.ox.ac.uk/ukbiobank/big40/] for Imaging Phenotypes GWAS.

## Acknowledgments

The authors thank the research participants contributing to the genome-wide association studies (GWASs) used for the present study, including the GWASs of Alzheimer’s disease and the GWASs of hippocampal volumes from UK Biobank. And this research was funded by the Dutch Research Council (ZonMW) VIDI no. 09150171910068; Amsterdam UMC Stimuleringsbeleid; and the Amsterdam Cohort Hub funded by the NWO ‘Sectorgelden’.

## Notes

### Competing Interest Statement

W.F. has been an invited speaker at Biogen MA Inc, Danone, Eisai, WebMD Neurology (Medscape), NovoNordisk, Springer Healthcare, European Brain Council. All funding is paid to her institution. W.F. is consultant to Oxford Health Policy Forum CIC, Roche, Biogen MA Inc, and Eisai. All funding is paid to her institution. W.F. participated in advisory boards of Biogen MA Inc, Roche, and Eli Lilly. W.F. is member of the steering committee of EVOKE/EVOKE+ (NovoNordisk). All funding is paid to her institution. W.F. is member of the steering committee of PAVE, and Think Brain Health. W.F. was associate editor of Alzheimer, Research & Therapy in 2020/2021. W.F. is associate editor at Brain.

