## Supplementary material for "Identification of novel associations of candidate loci with Alzheimer’s disease by leveraging the shared genetic basis with hippocampal volume": It contains required supplemenatry figures and tables.: Supplementary_Material.pdf

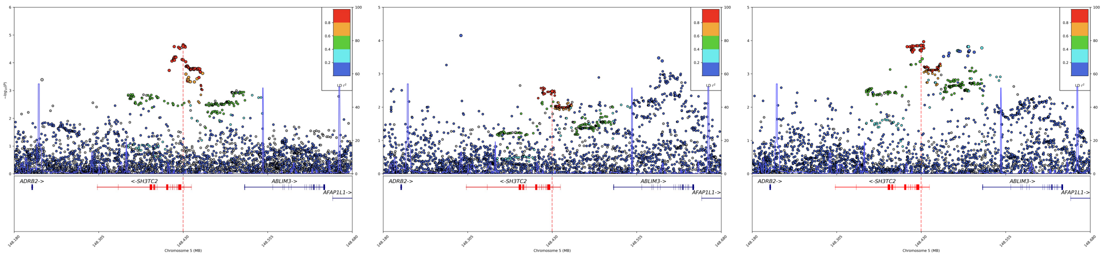

AD Left Hippocampus Right Hippocampus

Figure 1. regional plots of genetic variant rs36080, displayed as a locus zoom plot. The index SNP is annotated in purple, and nearby SNPs within 500KB ranges are colored according to how strong they are in LD (r2) with the index SNP, as derived from the 1000 Genomes LD reference. Left panel: regional plots based on AD GWAS; Middle panel: regional plots based on left hippocampal volume GWAS; Right panel: regional plots based on right hippocampal volume GWAS. AD: Alzheimer's disease; KB:Kilobyte; LD: linkage disequilibrium

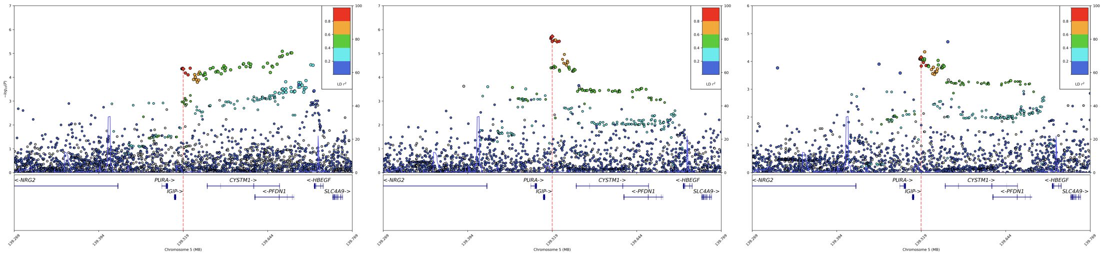

AD Left Hippocampus Right Hippocampus

Figure 2. regional plots of genetic variant rs156092

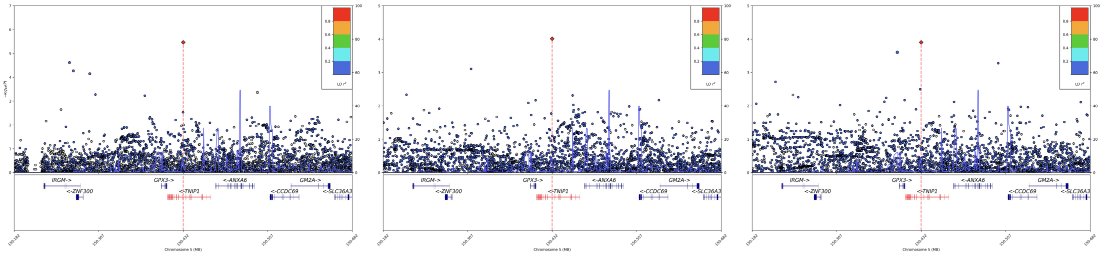

AD Left Hippocampus Right Hippocampus

Figure 3. regional plots of genetic variant rs871269

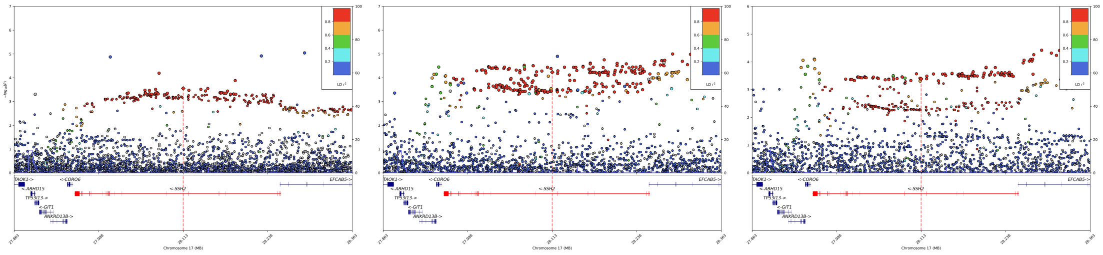

AD Left Hippocampus Right Hippocampus

Figure 4. regional plots of genetic variant rs2628166

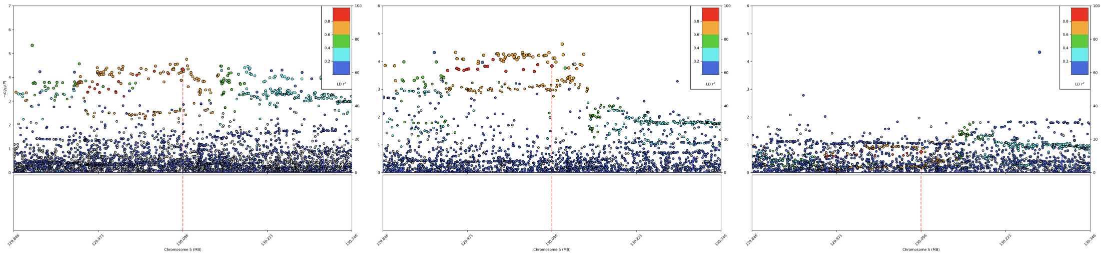

AD | Left Hippocampus | Right Hippocampus

Figure 5. regional plots of genetic variant rs12518350

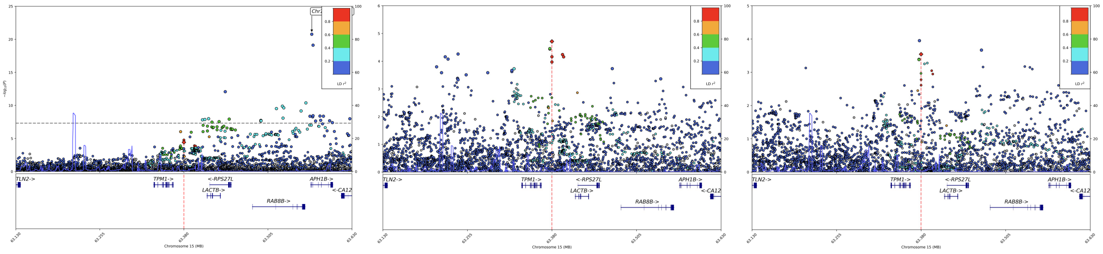

AD | Left Hippocampus | Right Hippocampus

Figure 6. regional plots of genetic variant rs12904537

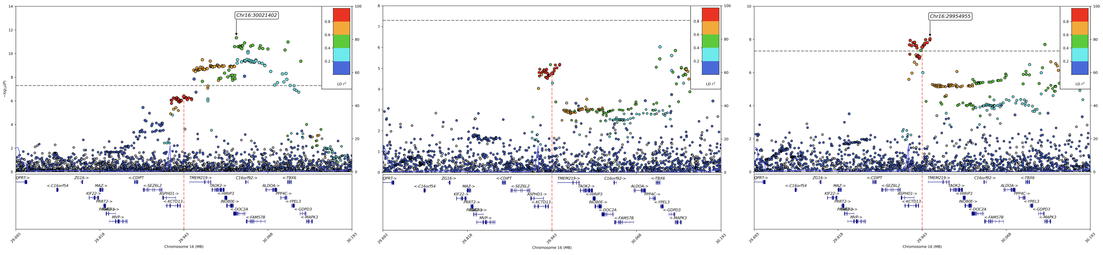

AD | Left Hippocampus | Right Hippocampus

Figure 7. regional plots of genetic variant rs12919683

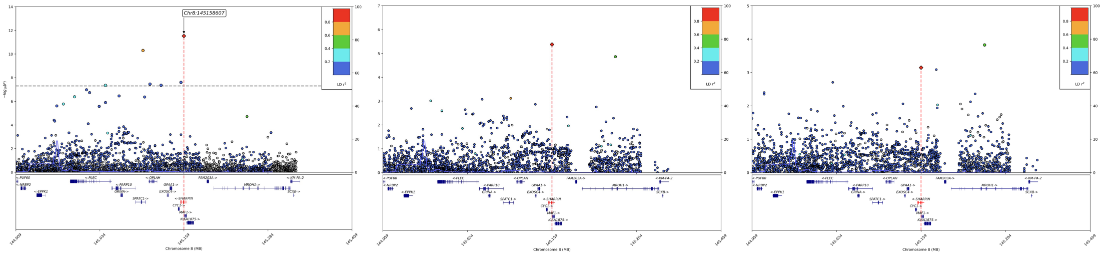


AD Left Hippocampus Right Hippocampus

Figure 8. regional plots of genetic variant rs34173062

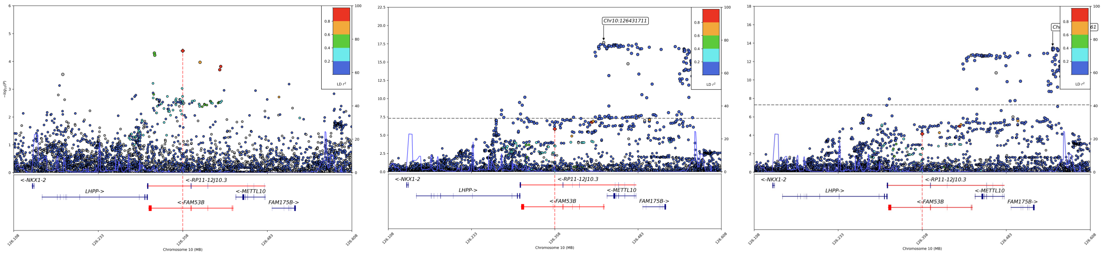


AD Left Hippocampus Right Hippocampus

Figure 9. regional plots of genetic variant rs61870529

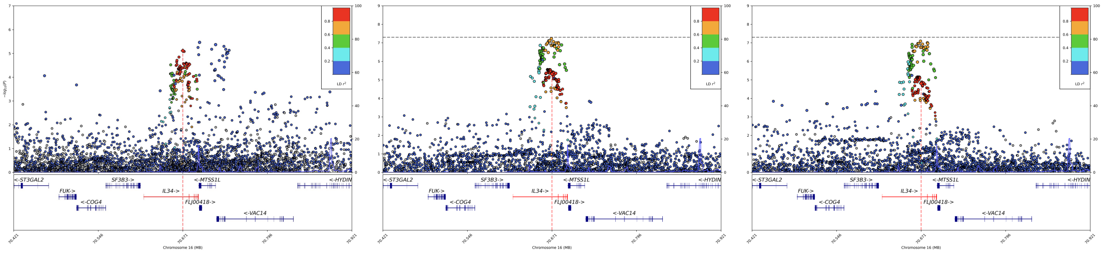


AD Left Hippocampus Right Hippocampus

Figure 10. regional plots of genetic variant rs139473334

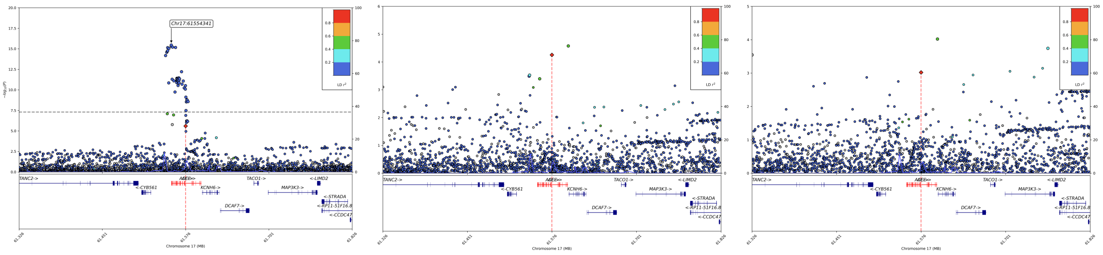


AD Left Hippocampus Right Hippocampus

Figure 11. regional plots of genetic variant rs149155892
